# Brief Report: An Eleven-Year Cessation Gap among Korean Former Smokers using Heated Tobacco or E-Cigarettes: Implications for Lung Cancer Risk Estimates

**DOI:** 10.64898/2026.08.11.26360188

**Authors:** S Cook, G Cohen, KM Cummings

## Abstract

**Introduction:** A Korean National Health Insurance Service (NHIS) cohort study reported higher lung cancer incidence among former smokers who used e-cigarettes after quitting than among those who quit without them. Interpreting that association requires knowing which product was used and how long ago participants quit; the cohort measured neither directly, but the contemporaneous Korea National Health and Nutrition Examination Survey (KNHANES) did.

**Methods:** KNHANES 2018, a cross-sectional national survey, used the NHIS umbrella e-cigarette term, a separate heated tobacco product (HTP) item, and months since cessation. Former smokers using either product (n = 43) were compared with those using neither (n = 1,305), with replication in 2019–2023.

**Results:** Of adults affirming current umbrella e-cigarette use, 64% also reported HTP use (former smokers, 50%); 79% of product-using former smokers reported HTP use. Median time since cessation was eight months among product users and twelve years among others (Kolmogorov–Smirnov D = 0.76, P < 0.001). The gap was replicated in every wave through 2023 (D = 0.63–0.76) and persisted in the long-term quitter stratum (pooled 2019–2023 medians, 6 vs 18 years), in Kim et al.’s high-risk subgroup (2 vs 12 years), after age matching (residual 9.0–11.0 years) and among umbrella-item respondents only (D = 0.73).

**Conclusions:** The reported association is consistent with residual confounding by cessation timing and with product misclassification. It also rests on few exposed events: 16 in the long-term stratum, and 31 of 71 retained when exposure was updated at the next screening. Because at most half of the exposure was exclusive liquid e-cigarette use, an e-cigarette-specific estimate would need about four times as many exposed events for the same precision. Repeated-screening designs that update cessation and product use at every screening, already used for cessation in the same database, would retain the full sample and allow more precise, product-specific dose-response estimates.

## Introduction

Kim Y.W. et al.^1^ recently analyzed a Korean National Health Insurance Service (NHIS) cohort and reported higher lung cancer risk among former smokers who used e-cigarettes after cessation than among those who quit without them. An accompanying commentary viewed these findings as evidence of long-term risks after switching to e-cigarettes.^2^ The study has several strengths, including a 4.5-million-participant national cohort and a switcher-versus-quitter comparison consistent with best practices for nonrandomized observational studies.^3^ However, attributing this association to post-cessation e-cigarette use requires sufficiently precise measurement of the product used, time since cigarette cessation, and duration of prospective use. Related concerns about reverse causation, product classification, continuity of use, and sparse exposed events have previously been raised qualitatively.^4^ Although the Korea National Health and Nutrition Examination Survey (KNHANES) cannot quantify reverse causation in the NHIS cohort, it can quantitate product use and cessation timing in the same national population and period. Because KNHANES has no lung cancer outcomes, our analysis is descriptive: it cannot reproduce or correct Kim et al.’s hazard ratios, but it can show what a comparably defined exposure group was likely to consist of. Using the 2018–2023 KNHANES, we show that half or more of a comparably defined exposure group used HTPs and that product users had quit about a decade more recently than other former smokers. In the Discussion, we consider what these findings, together with the absence of follow-up on product use, imply for the interpretation and precision of Kim et al.’s estimate. We close with a recommendation for a more precise and better powered approach following the methods of Park et al. in the same NHIS database.^5^

## Methods

We analyzed contemporaneous, nationally representative data from the 2018–2023 KNHANES to compare product use and time since cigarette cessation among product-using and nonproduct-using former smokers.^6–8^ We also examined whether differences in cessation duration persisted within strata corresponding to those used by Kim et al., within their high-risk subgroup (ages 50–80 years with ≥20 pack-years), after age matching, and when the exposed group was restricted to respondents reporting e-cigarette use. Because the wording of product items changed after 2018, we examined product composition using the 2018 survey, whereas cessation-duration analyses were replicated separately in each survey wave using year-specific product definitions; pooled analyses combined only the 2019– 2023 waves, which used the same items (see Supplement for detailed methods). Cessation-duration distributions were compared with two-sided Kolmogorov–Smirnov tests and the probability that a product user had quit more recently than another former smoker, and trends with age were estimated with Theil–Sen slopes; pack-years were calculated as cigarettes per day divided by 20, multiplied by years smoked. Analyses were unweighted; applying survey weights changed product shares by less than 2 percentage points. An age matching sensitivity analysis retained former smokers using neither product only if they were within 2 years of age of a product user (941 of 1,305 retained).

## Results

### Half to four-fifths of switchers were likely using heated tobacco products (HTPs)

Heated tobacco products (HTPs) were introduced in Korea shortly before the 2018 NHIS baseline and were widely used by 2018.^9,10^ The NHIS 2018 questionnaire used the umbrella e-cigarette term “전자담배” (jeon-ja-dam-bae) but did not have a separate HTP item, creating potential misclassification between liquid e-cigarettes and HTPs.^1,8^ KNHANES, fielded the same year, asked about use of the same umbrella e-cigarette term as NHIS, followed by a separate question about HTP use [Supplement, Section 2]. Among adults answering the first question affirmatively, 64% (110 of 171) also affirmed use of HTP [Supplement, Table S1]. Depending on how respondents interpreted the NHIS 2018 e-cigarette item, HTP users would be expected to comprise approximately 50%–79% of a comparably defined exposure group of former smokers [Supplement, Section 3]. Therefore, the reported association should not be interpreted as product-specific evidence about liquid e-cigarettes, as it has been.^1,2^

### Stratification narrowed but did not remove the cessation gap

A second concern is confounding by time since cessation. KNHANES 2018 measured time since cessation to the month and showed substantial separation between the groups. The median time since the last cigarette was eight months among HTP/EC-using former smokers and twelve years among those using neither product, with distributions that barely overlapped (Figure 1A: Kolmogorov–Smirnov D = 0.76, P < 0.001; HTP/EC users had quit more recently in 92% of pairwise comparisons). This pattern was reproduced in each KNHANES wave through 2023 (D = 0.63–0.76; all P < 0.001) [Supplement, Section 6]. This matters because the decline in lung cancer risk after cessation varies with time and population: in the same database, risk among men who had quit less than ten years earlier was similar to that of continuing smokers (aHR 0.98; for all cancers, 1.12),^5^ whereas among patients with COPD it fell within about five years (aHR 0.85).^11^ Time since cessation therefore needs to be measured precisely [Supplement, Section 7]; otherwise, an analysis comparing recent switchers with long-term quitters may attribute residual risk from recent cigarette smoking to post-cessation product use. Longer abstinence also selects people who did not relapse; recent quitters, including most switchers, remain at higher risk of relapse, which Kim et al. did not track.

**Figure 1.**
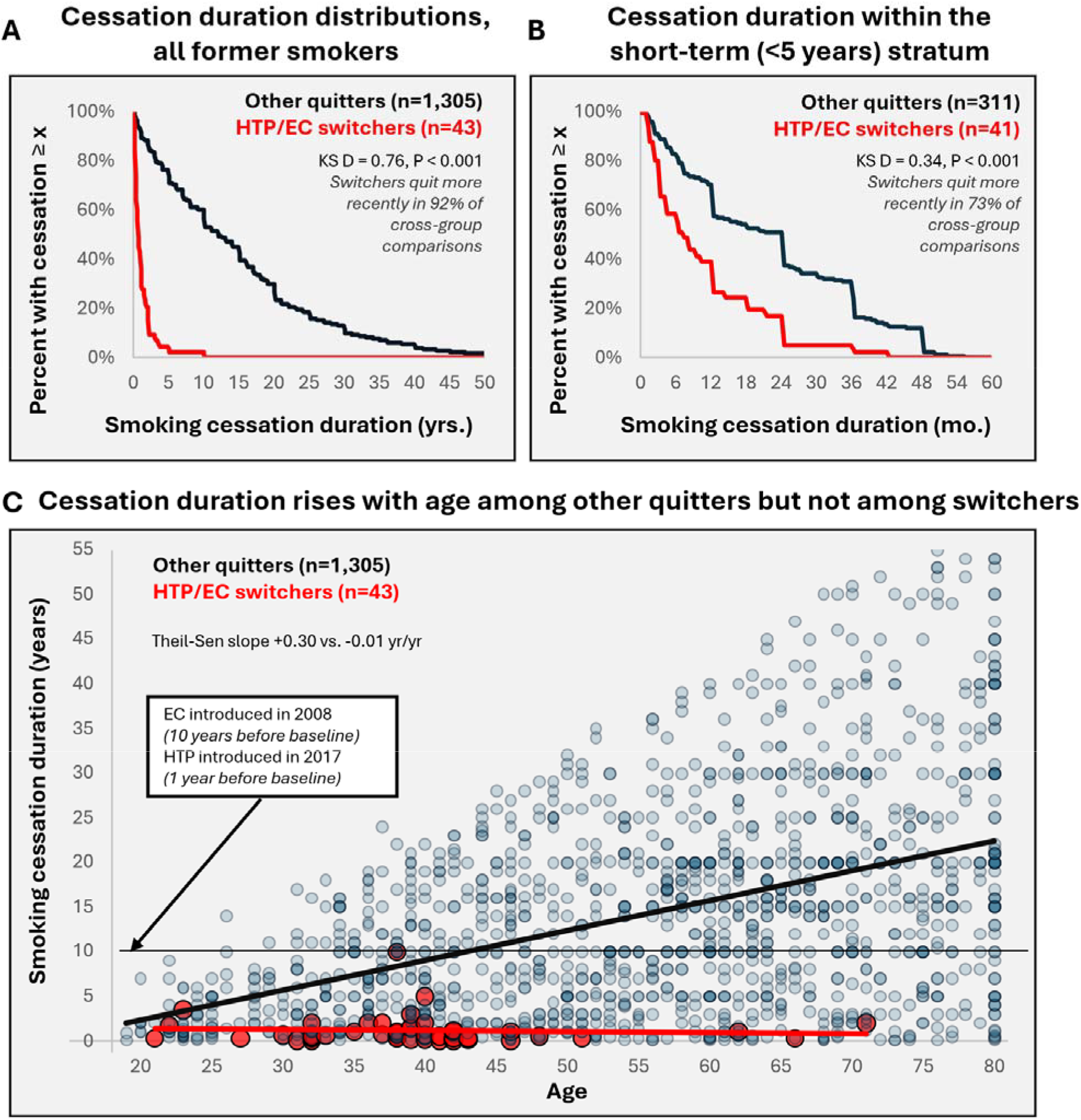
Smoking cessation duration among former smokers, KNHANES 2018. HTP/EC switchers (red; n = 43) vs. quitters by other means (blue; n = 1,305). (A) Complementary cumulative distribution across years for all former smokers (Kolmogorov–Smirnov D = 0.76; switchers quit more recently in 92% of cross-group comparisons; P < 0.001). (B) The same in months within the short-term (<5-year) stratum (D = 0.34, 73%, P < 0.001). (C) Age vs. cessation duration with per-group Theil–Sen fits and distribution-free 95% confidence intervals: +0.30 years of cessation per year of age (0.26 to 0.33) among quitters by other means and −0.01 (−0.04 to 0.01) among switchers, nonoverlapping. The dashed line indicates cessation beginning in 2008, around when e-cigarettes first went on sale in Korea; HTPs reached the market in June 2017.

Kim et al. addressed cessation timing by stratification, but the strata only crudely captured cessation duration. Their short-term and long-term quitter categories were derived from smoking status at two screening waves rather than exact quit dates; “long-term” quitters were already former smokers at the 2012–2014 screening, with no upper limit on cessation duration. In KNHANES, stratification narrowed but did not eliminate the imbalance. In 2018, within the short-term quitter stratum, the cessation-duration distributions remained separated (Figure 1B: Kolmogorov–Smirnov D = 0.34, P < 0.001), and HTP/EC-using former smokers had quit more recently in 73% of pairwise comparisons; in later waves this difference narrowed (0.5 years in 2019) and was absent from 2020 onward. The imbalance that persisted was in the long-term stratum: pooling the 2019–2023 waves, which used the same product items, median cessation duration was 6.0 years among 135 product users versus 18.0 among 4,942 former smokers using neither product (D = 0.66, P < 0.001) [Supplement, Section 6].

### Age adjustment and high-risk restriction do not remove the imbalance in quit duration

Age adjustment is also an incomplete proxy for cessation duration. Among former smokers not using either product, older age was associated with more time since quitting; among those with HTP/EC use, cessation was recent across the observed range of ages (Figure 1C). The Theil–Sen slope of cessation duration by age was +0.30 years per year among no-product quitters (95% CI 0.26 to 0.33) and −0.01 among product users (−0.04 to 0.01); the intervals did not overlap. After age matching, a 9.3-year difference remained in 2018, with differences of 9.0–11.0 years across the six survey waves [Supplement, Section 6]. These differences also persisted when the exposed group was restricted to respondents affirming the umbrella e-cigarette item (D = 0.73) [Supplement, Table S3]. The gap also persisted in Kim et al.’s high-risk subgroup (ages 50–80 years, ≥20 pack-years), which yielded their largest incidence estimate (aHR 1.91). Pooling 2019–2023, median cessation duration in this subgroup was 2.0 years among 63 product users versus 12.0 among 1,906 former smokers using neither product (D = 0.60, P < 0.001), with slightly lower mean pack-years among product users (34 vs 38) [Supplement, Section 8]. Eligibility criteria select on age and cumulative exposure; they do not balance time since cessation.

## Discussion

### A 4.5-fold shift on adjustment highlights the importance of precise quantification of covariates

In Kim et al.’s cohort, switchers had crude lung cancer incidence-rate ratios of 0.29 and 0.39 relative to short-term and long-term non-product using quitters, respectively. Adjustment for the modeled covariates yielded adjusted hazard ratios of 1.31 (95% CI, 1.00–1.71) and 1.74 (95% CI, 1.07–2.84) respectively; the findings depend on the precision of an adjustment which reversed the direction of the association and shifted the estimates approximately 4.5-fold. Such a large reversal can result from appropriate adjustment for confounding, but here it relies on age and two-wave quitter categories standing in for time since cessation, on exposure that was not reassessed during follow-up, and on a small number of exposed events [Supplement, Section 9].

### Evaluating persistence of switching vs. quitting at 1–2 years diminished all effect sizes

Prospective product use, switching, and relapse were not reassessed, with one exception. In time-varying models restricted to the 77% of the cohort rescreened in 2019–2020, exposure was updated when participants changed state, and all estimates decreased: the lung cancer incidence moved from 1.56 (95% CI 1.24–1.97) to 1.38 (0.98–1.98) in the overall cohort and from 1.91 (1.44–2.53) to 1.40 (0.83–2.37) in the high-risk subgroup [Supplement, Section 10]. With a single reassessment, both incidence estimates lost statistical significance, and the excess risk in the high-risk subgroup fell by more than half (aHR 1.91 to 1.40).

### The long-term quitter contrast rested on 16 lung cancer events and was subject to previously described confounds

Despite a cohort of 4.5 million, precision depends on the number of exposed events, and two features reduce what those events can show about e-cigarettes. The long-term contrast rested on 16 exposed lung cancer events, adjusted for ten covariates. Liquid e-cigarettes made up at most half, and possibly one-fifth, of a comparably defined exposure [Supplement, Section 3]; an e-cigarette-specific estimate would need about four times as many exposed events for the same precision [Supplement, Section 11]. In addition, when exposure was updated at the next screening, 31 of 71 exposed incidence events and 3 of 19 lung cancer deaths remained. Kim T. et al. lost exposed participants in the same way when it checked whether switching persisted.^11^ This loss follows from classifying exposure once and setting aside people whose behavior changed.

### The dual-use comparison was the most precisely estimated but was also subject to confounds

The comparison least affected by cessation timing appears only in the supplement: dual users against cigarette-only smokers, with 578 exposed lung cancer events (aHR 0.78, 95% CI 0.72–0.85; 0.94, 0.83– 1.07, in the high-risk subgroup), 114 lung cancer deaths (0.66, 0.55–0.79) and 1,459 deaths from any cause (0.74, 0.71–0.78).^1^ Both groups were current smokers, so cessation timing could not differ between them. We draw no causal inference from it, since product mix, smoking intensity, and persistence of dual use were not reported, but it is the most precisely estimated product comparison in the paper.

### Limitations

While our analysis of the contemporaneous national KNHANES data allows us to assess data not measured in the Kim et al. study, several limitations are important to note. First, KNHANES and NHIS include different respondents, so KNHANES cannot directly establish product composition or cessation histories in Kim et al.’s cohort. Rather, the KNHANES analysis allows us to characterize comparably defined groups in the same national population and period. Second, product use and smoking histories were self-reported and product item specificity changed across survey waves. Third, KNHANES cannot examine lung cancer incidence prospectively, directly examine the associations observed by Kim et al., or test for reverse causation.

### Opportunities for more precise and accurate future studies

None of these findings show that switching to e-cigarettes or HTPs is safe, and we do not claim that it does. Rather, Kim et al. lack the measurement resolution needed to determine whether post-cessation use of e-cigarettes or HTPs affects lung cancer risk relative to quitting without these products. However, the same national data support a design that keeps the full sample: updating smoking and product status at every biennial screening, so that relapse is modeled rather than excluded and both time since quitting and duration of product use accumulate as exposure. Park et al. have demonstrated this approach to cessation in NHIS [Supplement, Section 12].^5^ A similar methodology, applied to products, with the separate HTP and liquid e-cigarette items available from 2019, would give more precise dose-response estimates for each product. Until such studies are performed, the association reported by Kim et al. should be treated as a signal requiring replication rather than as definitive evidence that post-cessation e-cigarette and/or HTP use independently increases lung cancer risk.

## Acknowledgments

A preprint of this study was posted on the MedRxiv preprint server.^12^

## Data availability

KNHANES public use data is available at https://knhanes.kdca.go.kr/knhanes/main.do.

## Author contributions

All authors were involved in writing and review. Data analysis was performed by G.C. and S.C.

## Disclosure of computational tooling

Preliminary analysis of the KNHANES public-release files was performed with the assistance of Claude (Anthropic). The authors generated the figures and verified the descriptive statistics, distributional comparisons and rank tests using Excel.

