## Supplement I for "Brief Report: An Eleven-Year Cessation Gap among Korean Former Smokers using Heated Tobacco or E-Cigarettes: Implications for Lung Cancer Risk Estimates"

### Preface

The study of Kim et al. was designed to provide insights into the impact of electronic cigarettes in Korea, using the NHIS database. Here, we evaluate the assessment precision of the study, with particular focus on understanding the time quit of each of the cohorts and product mix used.

### 1. Methods

**Data source.** All analyses in this appendix use the public-release microdata of the Korea National Health and Nutrition Examination Survey (KNHANES) from 2018 to 2023 [7]. The KNHANES is a nationally representative, annually fielded cross-sectional survey conducted by the Korea Disease Control and Prevention Agency [6], drawing on the same adult population and the same calendar years as the National Health Insurance Service (NHIS) screening cohort analyzed by Kim et al. [1]. Kim et al. neither analyze nor cite KNHANES; we use it because it is the national reference source for tobacco use surveillance in Korea. In Kim's 2018 baseline year, it records two quantities which the NHIS screening item does not (time quit smoking and product used). The 2018 NHIS screening questionnaire asked a single question about "e-cigarette" (전자담배) use, with responses coded as days of use in the previous month (none, 1 to 2, 3 to 9, 10 to 29, or every day). The questionnaire did not use the more specific "liquid nicotine-containing e-cigarette" terminology and had no separate heated tobacco item [8]; it could neither separate the products nor date the quit point.

**Population and definitions.** Analysis is restricted to adults aged 19 years and older. The smoking-status item is administered only to respondents who have smoked, so BS3_1 = 3 identifies former smokers without a further lifetime-use filter. Current smokers are BS3_1 in{1 (daily), 2 (occasional)}; former smokers are BS3_1 = 3. Cessation duration is BS6_4, recorded in months and divided by 12, excluding codes of 8888 and above, with at least 30 days of abstinence required. Current HTP/EC use is the union of the EC item and the HTP item. The HTP/EC group comprises former smokers currently using either product; the no-product group comprises former smokers using neither. Short-term and long-term strata are defined at 5 years of cessation (short-term less than 5 years) to mirror the two-screening classifier described by Kim et al.

**Sample size and exclusions.** For the 2018 baseline, the flow is 7,992 total records, then 6,489 adults aged 19 and older, then 1,356 former smokers (BS3_1 = 3), then 1,355 with a valid cessation duration, and then 1,348 with at least 30 days of abstinence, of whom 43 were current HTP/EC users and 1,305 used neither. Two exclusions apply: one former smoker with a refusal or missing cessation code (BS6_4 at least 9000) and seven reporting fewer than 30 days of abstinence (none of whom were HTP/EC users). In every wave, the 30-day rule excludes fewer than ten respondents (7, 5, 5, 2, 0 and 2 in 2018 to 2023) and leaves every reported median unchanged. The per-wave and per-group exact n values are listed in Tables S1, S3 and S6 and in Figures 1 and S3.

**Weighting and external benchmark.** The comparisons reported below are internal relative comparisons (product composition, cessation-duration contrasts, and age-matched residuals) that do not depend on population projection and are therefore computed unweighted on the analytic sample. Where an absolute national prevalence is at issue, the survey design must be applied, and we benchmark against a peer-reviewed KNHANES analysis. Applying the KNHANES sampling weights to the 2019 file, the current HTP use under the standard definition (daily or occasional) is 8.6% among adult men, matching the 8.8% reported by Lee et al. [10] for the same wave using the same definition; female HTP use is 1.5% in both. The exposure rule used by Kim et al., which counts only daily use, yields 5.4% among men against the same weights, undercounting the published national estimate by approximately one-third. Weighted male liquid-EC use in 2019 was 4.2%, which is less than male HTP use, which is consistent with HTP being the predominant novel product in the exposed group. These weighted figures are reported only to establish that the corrected HTP definition reproduces the national benchmark and that the daily use restriction departs from it; the within-sample contrasts elsewhere in this appendix are unweighted, as noted, and do not depend on the benchmark.

**Statistical methods.** Cessation duration is strongly right skewed and bounded at zero, so normality and equal variance are not assumed anywhere: the median and interquartile range (IQR) summarize it, and distributions (Figure 1A and 1B) are compared with the two-sample Kolmogorov–Smirnov test, the two-sided Mann–Whitney rank-sum test, and the probability of superiority (the proportion of cross-group pairs in which the HTP/EC switcher had quit more recently, ties counted as one half). In 2018, the median [IQR] cessation duration was 0.7 (0.25–1.5) years among HTP/EC switchers versus 12.0 (5.0–20.0) years among no-product quitters; means are shown in Table S2 only for comparability with Kim et al. The age-matched comparison restricts the no-product group to former smokers for whom at least one HTP/EC switcher falls within 2 years of age (n = 941 of 1,305 in 2018); the residual gap is the median cessation of the matched pool minus the median among HTP/EC switchers. In Figure 1C, the age–duration relationship is summarized per group by Theil–Sen robust regression with distribution-free 95% confidence intervals and a Kendall rank correlation; non-overlap of the intervals is a conservative sufficient condition for a difference in slopes rather than a direct test of it. As a parametric check, an ordinary-least-squares age-by-group interaction is estimated with a heteroskedasticity-consistent (HC1) covariance matrix, because the dispersion of cessation duration differs by group by construction (standard deviation 12.11 years among no-product quitters and 1.72 years among product users) and a pooled-variance model would assign the comparator's dispersion to the exposed group; pooled-variance values are given alongside for transparency. Our models contain age, group and their interaction and no other covariates; no covariate screening or selection was performed. The dashed reference line in Figure 1C marks a cessation duration of 10 years, the interval between the 2008 introduction of e-cigarettes in Korea and the 2018 baseline. All tests are two-sided and unweighted; exact P values are reported where estimable and as P < 0.001 otherwise. The same measures are computed separately in each wave from 2018 to 2023 using year-specific product definitions, and pooled across the 2019 to 2023 waves, which used the same product items (§6 to §8); 2018 is always reported separately because its items differed. Respondents differ between waves, and the pooled figures are unweighted.

**Survey design**. KNHANES is a stratified multistage cluster sample. The internal contrasts are computed unweighted and without a clustering adjustment, consistent with the internal-comparison framing above; the weighted prevalence benchmark above is the quantity for which the design is applied. All observations are distinct respondents measured once; no respondent is measured repeatedly.

**Test statistics and effect sizes.** For the 2018 overall contrast, Mann-Whitney U = 4,391 (n1 = 43, n2 = 1,305; P = 4.2 x 10-21) and Kolmogorov-Smirnov D = 0.758 (P = 3.3 x 10-25). Within the short-term stratum, U = 3,480 (n1 = 41, n2 = 311; P = 2.1 x 10-6) and D = 0.341 (P = 0.0003). The age-by-group interaction coefficient is -0.359 (HC1 SE 0.024; 95% CI -0.407 to -0.312); the pooled-variance standard error for the same coefficient is 0.166 (95% CI -0.686 to -0.033; df = 1,344), which is the value a homoskedastic model returns when the comparator's 12.11-year dispersion is assigned to an exposed group whose own dispersion is 1.72 years. The Theil–Sen slope of cessation duration on age is +0.296 years per year among no-product quitters (95% CI 0.259 to 0.333; Kendall tau = +0.309, P = 4.1 x 10-60) and -0.008 among product users (95% CI -0.042 to 0.012; tau = -0.090, P = 0.42), a difference of -0.304. The two intervals do not overlap, and the clear space between them, 0.247 years per year of age, is about three to five times the width of either interval. These are the estimates quoted in the main text and shown in Figure 1C. Under ordinary-least-squares fits to the same data the slopes are +0.349 (SE 0.019) and -0.011 (SE 0.027), a difference of -0.359; combining the two group-specific standard errors gives SE 0.033 and a 95% CI of -0.424 to -0.295, and the heteroskedasticity-consistent values are SE 0.024 and -0.407 to -0.311. The least-squares and Theil–Sen estimates agree to within 0.06 years per year of age; the Theil–Sen value is preferred because cessation duration is right skewed and bounded at zero, as set out above. Effect sizes are the Kolmogorov-Smirnov D, the probability of superiority computed as U divided by the product of the two group sizes with ties counted as one half, the Theil-Sen slopes, and the age-matched residual gap in years. No Bayesian analysis was performed.

**Software and reproducibility.** Analyses were initially performed in Python and statsmodels (interaction model) and then manually reverified in Excel. EC and HTP definitions and derivations are shown in Figures S1 and S2.

**Data and code availability.** The KNHANES public-release microdata (2018 to 2023) are freely available from the Korea Disease Control and Prevention Agency at https://knhanes.kdca.go.kr.

### 2. Questionnaire items for e-cigarettes and heated tobacco products

The KNHANES 2018 questionnaire included a first question asking about current e-cigarette use (questionnaire item Q11-1; variable BS12_2; see Figure S1). Here, the term e-cigarette was similarly generic and not specified to be “liquid nicotine-containing”. It was then followed by a checklist (questionnaire item Q12-1; heated tobacco option, variable BS12_47) asking whether certain products were currently used, excluding e-cigarettes, and one of the choices was heated tobacco products, defined with specific examples (IQOS, glo). Therefore, we can deduce that someone who responded affirmatively to question 12 but negatively to question 11 was exclusively using HTP. Likewise, yes to 11 and no to 12 would correspond to exclusive liquid EC use. Yes to both is ambiguous and could mean either the dual use of both or the exclusive use of HTP. See Figure S1 for the questions asked in Korean.

From KNHANES 2019 onward, the questions are asked more specifically (see Figure S2). Question 4-1 asks about current HTP use and provides specific examples. This is followed by question 5-1, which asks about liquid nicotine EC use (see Figure S2). From 2019, current HTP use is defined as daily or occasional use (BS12_47 = 1 or 2) and current liquid EC use as BS12_2 = 1; in 2018, current HTP use is the checklist item (BS12_47 = 1) and current EC use the umbrella item (BS12_2 = 1).

Cessation duration was recorded in months (BS6_4, in every wave), whereas the Kim study captured only smoking status at two screening dates five years apart. From 2019, the KNHANES standardized the HTP question into daily, occasional, and former categories with frequency of use also asked, refining the product distinction it already drew in 2018 rather than introducing one.

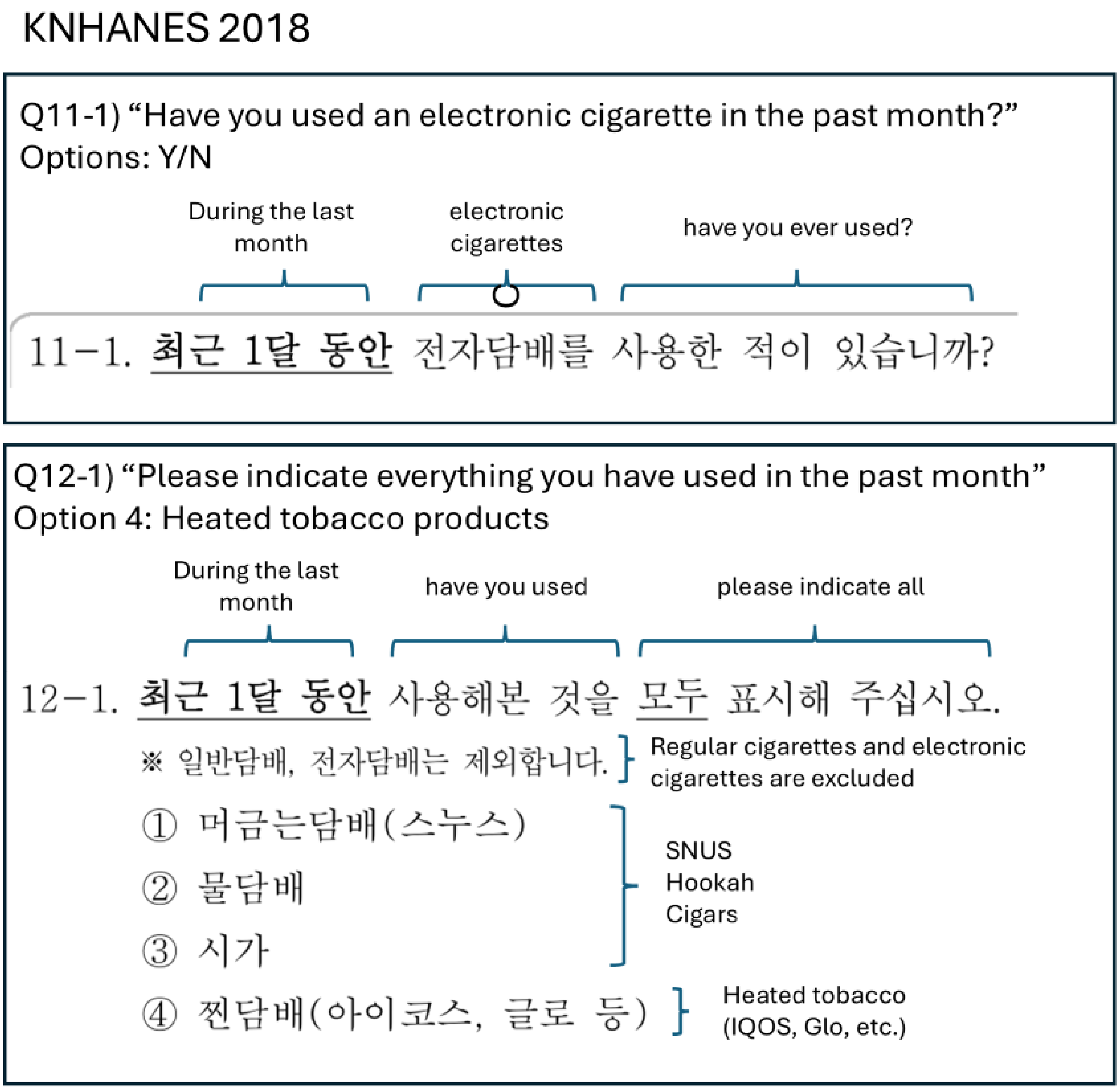


**Figure S1.** E-cigarette and HTP current use questions, KNHANES 2018.

*In 2018, participants were first asked about e-cigarette use, followed by a question on heated tobacco products. The e-cigarette term did not specify liquid nicotine, while the HTP included specific brands as examples (IQOS, glo).*

### KNHANES 2019 onward

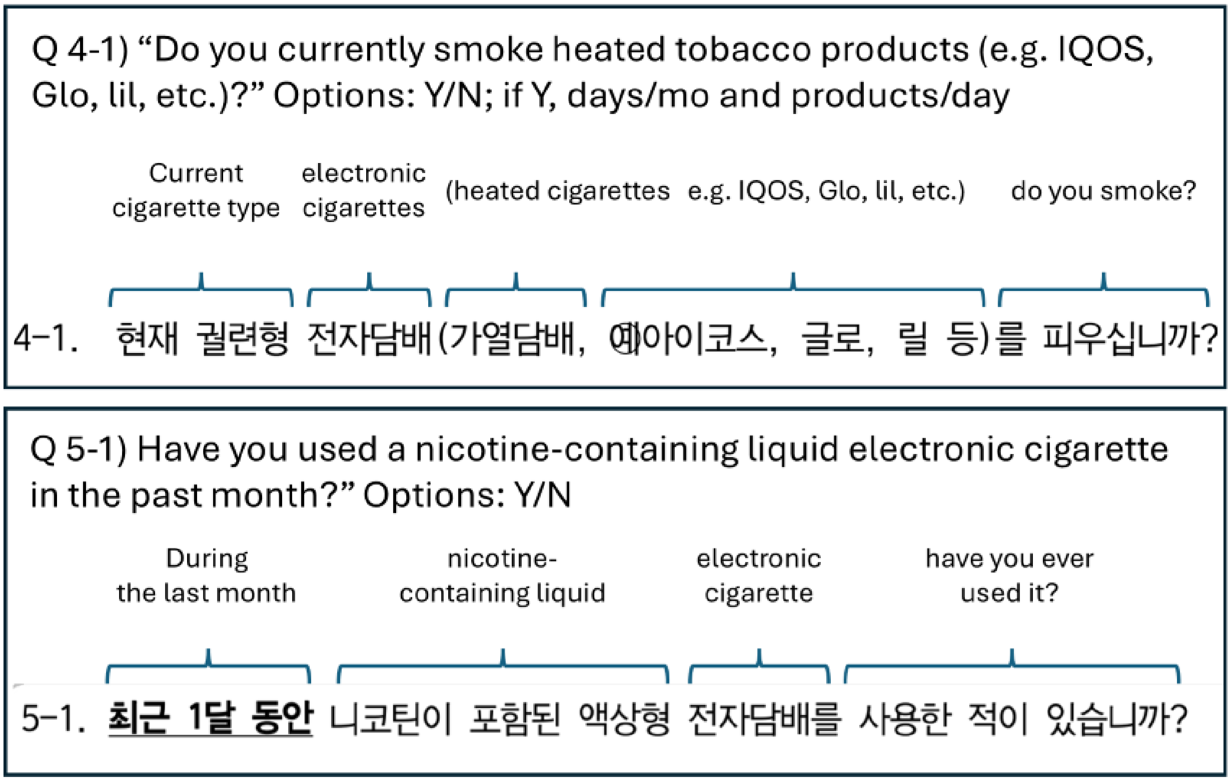


**Figure S2.** E-cigarette and HTP current use questions, KNHANES 2019 onward.

*In 2019, participants were first asked about HTP use, followed by a question on nicotine-containing liquid e-cigarettes. The e-cigarette term specified nicotine-containing liquid EC, and the HTP included specific brands as examples (IQOS, Glo, lil).*

### 3. Product composition

**Product classification.** The NHIS 2018 item used only the umbrella term 전자담배 and had no heated tobacco item [8], so a respondent using an HTP had no other place to report it. The authors themselves note that other publications describe the same questionnaire-derived exposure by a broader umbrella term, citing Choi et al. [13] (their reference 47), whose account of the identical item states that it does not differentiate heated tobacco products from nicotine vapor products. An independent KNHANES analysis agrees: Lee et al. [10] caution that some 2018 cigarette-and-HTP dual users may have been misclassified as cigarette-and-EC dual users. In the raw 2018 file, 205 respondents reported current HTP use on the explicit HTP checklist, but only 171 reported current use on the umbrella "e-cigarette" item; only 110 of the 205 HTP users (54%) also affirmed the umbrella item, so an umbrella item of the kind the NHIS used would have missed 95 HTP users (46%) rather than subsuming them. Because the NHIS item did not distinguish products, the exposed group's product mix can be bounded but not measured, and the two natural readings of a single e-cigarette question bound it narrowly. If the item captured only respondents who identified with the narrow e-cigarette term, as KNHANES's separate 2018 e-cigarette item did, the exposed group would comprise only those responders: of the 43 KNHANES switchers, 18, of whom 9 (50%) also used HTP, the 25 HTP-only switchers being misclassified into the comparator. If instead the item did not differentiate HTP from vapor, as the authors' own cited source (Choi et al.) describes, it would capture all product users, of whom 79% used HTPs in KNHANES (Table S1, Panel B). Across all adults, 110 of the 171 respondents (64%) who affirmed the umbrella item also reported HTP use (Table S1, Panel C). The former-smoker share (9 of 18, 50%) is the lowest of the smoking-status groups (most represent dual-users with cigarettes). The HTP use in the exposed group therefore falls between 50% and 79% under these two readings. An HTP frequency item exists only from 2019; thus, a daily use restriction is not definable for HTP/EC in 2018.

**Table S1.** Exposure composition among HTP/EC users, KNHANES 2018 and 2019

| Metric | KNHANES 2018 | KNHANES 2019 |
|---|---|---|
| ***A. Combustible-cigarette status among ever-smokers currently using HTP or EC*** | | |
| **Dual users (current combustible + HTP/EC), n** | **222** | **214** |
| **Complete switchers (former combustible + HTP/EC), n** | **43** | **101** |
| Total, n | 265 | 315 |
| Complete switchers, % | 16.2% | 32.1% |
| **Dual-user : switcher ratio** | **5.2 : 1** | **2.1 : 1** |
| ***B. Product type among HTP/EC-using former smokers, n (%)*** | | |
| **HTP only** | **25 (58%)** | **62 (61%)** |
| **HTP, liquid-EC status unconfirmed** | **9 (21%)** | **N/A** |
| **HTP and liquid EC (confirmed dual)** | **N/A** | **14 (14%)** |
| **Liquid EC only** | **9 (21%)** | **25 (25%)** |
| Total HTP/EC former smokers, n | 43 | 101 |
| ***C. Umbrella e-cigarette item among all adults, by smoking status*** | | |
| Current smokers affirming the item: also HTP, n/N (%) | 100/152 (66%) | N/A |

| Metric | KNHANES 2018 | KNHANES 2019 |
|---|---|---|
| Former smokers affirming the item: also HTP, n/N (%) | 9/18 (50%) | N/A |
| Never smokers affirming the item: also HTP, n/N | 1/1 | N/A |
| All adults affirming the item: also HTP, n/N (%) | 110/171 (64%) | N/A |
| Any HTP among all adult HTP/EC users, n/N (%) | 205/266 (77%) | 244/317 (77%) |
| ***D. Use frequency among current HTP users*** | | |
| Former-smoker HTP users: daily, n (%) | not measured | 67 (88%) |
| Former-smoker HTP users: occasional, n (%) | not measured | 9 (12%) |

*Panel A: switchers are the analytic sample (former smokers with a valid cessation duration of at least 30 days); dual users are current smokers reporting HTP or EC use for whom no cessation filter applies. Panel B denominator: HTP/EC-using former smokers in that analytic sample. In 2018, the e-cigarette item was an undifferentiated umbrella, so a positive response alongside HTP confirms HTP use but not liquid-EC use; that cell is therefore reported as HTP with unconfirmed liquid-EC status, and confirmed dual use is not definable for 2018 (N/A). The 2019 e-cigarette item was liquid specific, so its dual-use cell is genuine. In contrast to 2019, a share of the 2018 unconfirmed cells are likely exclusive-HTP umbrella responders. Panel C: adults aged 19 years or older of any smoking status, KNHANES 2018 only, because the 2019 e-cigarette item was liquid specific; the any-HTP row is shown for both years. Panel D: Among former-smoker HTP users in 2019, occasional users report a median of 10 days of use per month (IQR 3 to 10) within the 10-to-29-day band of the exposure question; among all adult current HTP users, 37% are occasional and are therefore counted as nonusers under a daily use exposure definition. No HTP frequency item existed in 2018; thus, daily use status is not definable for that wave. Percentages may not sum to 100 owing to rounding. Interquartile ranges for 2018 (years) are 0.25 to 1.50 (HTP/EC, all) and 5.00 to 20.00 (no-product, all); within the short-term stratum, 0.25 to 1.17 and 0.62 to 3.00; within the long-term stratum, 6.25 to 8.75 (n = 2) and 10.00 to 24.00.*

### 4. Cessation duration in 2018: the dominant confound

The two groups differ in how long ago they quit, and the difference does not go away when we account for it. Figure 1 (Panel A) shows the overall contrast (median 0.7 vs 12 years in 2018), Panel B shows that in 2018 it persisted within the short-term stratum used by Kim et al., and Panel C shows that it survives exact age matching. The overall contrast, the long-term stratum contrast and the age-matched residual are reproduced in every wave from 2018 to 2023 (§6). The short-term difference is specific to the early waves: it narrows from 1.4 years in 2018 to 0.5 years in 2019 and is absent from 2020 onward, so the imbalance that bears on Kim et al.'s estimates is concentrated in the long-term stratum (§7).

**Table S2.** Cessation duration among former smokers by product use group, KNHANES 2018 and 2019

| Metric | 2018 | 2019 |
|---|---|---|
| ***A. Cessation duration, all former smokers (years)*** | | |
| **HTP/EC group: median** | **0.7** | 2.0 |
| HTP/EC group: mean | 1.2 | 2.0 |
| HTP/EC group: n | 43 | 101 |
| **No-product group: median** | **12.0** | 12.0 |

| Metric | 2018 | 2019 |
|---|---|---|
| No-product group: mean | 14.5 | 14.7 |
| No-product group: n | 1,305 | 1,326 |
| ***B. Within-stratum cessation duration, median (years); n in parentheses*** | | |
| **Short-term (<5 yr): HTP/EC** | **0.6 (41)** | 1.5 (92) |
| **Short-term (<5 yr): no-product** | **2.0 (311)** | 2.0 (312) |
| **Long-term (>=5 yr): HTP/EC** | **7.5 (2)** | 6.0 (9) |
| **Long-term (>=5 yr): no-product** | **16.0 (994)** | 16.0 (1,014) |
| ***C. Residual after exact age-matching (+/- 2 yr)*** | | |
| HTP/EC: median age (yr) | 39 | 40 |
| Age-matched no-product: median cessation (yr) | 10.0 | 11.0 |
| **Residual cessation gap (yr)** | **9.3** | 9.0 |
| Maximum cessation duration among HTP/EC switchers (yr) | 10.0 | 9.0 |
| No-product quitters exceeding that maximum, % | 53.3% | 60.9% |

*HTP/EC group = former smokers currently using HTP or liquid EC; no-product group = former smokers using neither. The panel B stratum boundary is 5 years (short-term less than 5). Panel C: the no-product group is restricted to former smokers for whom at least one HTP/EC user falls within 2 years of age (n = 941 of 1,305 in 2018); the residual is the median cessation of that matched pool minus the median among all HTP/EC users. In 2018, the longest-abstinent HTP/EC switcher had quit 10.0 years earlier; in 2008, the year the authors gave for the arrival of e-cigarettes in Korea, 696 of 1,305 no-product quitters (53.3%), and 696 of 994 within the long-term stratum (70.0%), had quit longer ago than any switcher in the sample. The 2018 long-term HTP/EC median rests on two respondents and is reported for completeness only; it should not be read as a stable estimate. The long-term contrast is more securely read from 2019, when the HTP/EC long-term cell contains nine respondents and the same ordering holds (6.0 vs 16.0 years), and from the pooled 2019 to 2023 waves (135 respondents; §6).*

**5. The cessation-duration gap holds within a product-use population restricted to users affirmatively indicating that they used “전자담배” (electronic cigarettes, broad term)**

The product composition of the NHIS-exposed group cannot be measured and is only bounded: on the two natural readings of a single e-cigarette question, HTP users make up between 50% and 79% of it (§3). Section 4 reports the cessation-duration contrast for the full product-using group, in which HTP users constitute the majority. This section asks whether that contrast depends on them. It does not. Under the narrowest reading, in which every exposed respondent affirmed the e-cigarette item, half of them also affirmed using an HTP. The median exposed respondent had quit six and a half months before the survey against twelve years in the comparator, and the separation in Table S2 is essentially preserved.

**Definition of the restricted group.** Of the 43 product-using former smokers in KNHANES 2018, 18 answered the umbrella e-cigarette item (BS12_2 = 1); 9 of those 18 also reported HTP use on the separate checklist, and 9 reported liquid EC alone. The remaining 25 were HTP-only, identified by the checklist but not by the e-cigarette item. Restricting the exposed group to the 18 reproduces the narrow reading, under which every exposed respondent is an e-cigarette user by their own identification.

**Results.** The restricted definitions are compared against the full group in Table S3. The overall separation is essentially unchanged (D = 0.73 against 0.76; in both, the exposed respondent had quit more recently in 92% of cross-group comparisons). Within the short-term stratum, the separation is slightly wider (D = 0.37 against 0.34), and exact age matching leaves a residual gap of 9.5 years against 9.3 (P < 0.001 in both). The ceiling tightens: The longest-abstinent e-cigarette-item responders had quit 5.0 years before the survey, and 926 of 995 long-term comparators (93.1%) had quit longer ago than any exposed respondent did, compared with 70.0% in the full group. Within the 18, the 9 reporting liquid EC alone had a median cessation of 1.0 years, and the 9 also reporting HTP 0.3 years; both rest on nine respondents and are reported for completeness rather than as stable estimates.

**What the restriction costs is precision, not effect size.** Every measure of the size of the separation is essentially preserved: D moves from 0.76 to 0.73 overall and from 0.34 to 0.37 within the short-term stratum, and the proportion of cross-group comparisons in which the exposed respondent had quit more recently is unchanged at 92% and 73%, respectively; the age-matched residual widens from 9.3 to 9.5 years, and the age-by-group interaction coefficient is unchanged at -0.36. The rank-test P values move with the number of exposed respondents rather than with the effect: in the short-term stratum the number decreases from 41 to 17, and the Mann-Whitney P increases from less than 0.001 to 0.002 for an identical 73%. The interaction behaves differently, and the apparent loss of precision under restriction is an artifact of the variance assumption rather than of sample size. Under a pooled-variance model the standard error rises from 0.17 to 0.23 and P from 0.03 to 0.12; under the heteroskedasticity-consistent estimator used throughout (§1), the robust standard error is 0.024 in the full group and 0.023 under both restricted definitions, and the coefficient is -0.359, -0.362 and -0.355, respectively. Restriction therefore leaves the interaction essentially unchanged in both estimate and precision. The reason is that the exposed group's dispersion falls further under restriction, from 1.72 to 1.30 years, so a pooled-variance model penalizes it more heavily even as the underlying contrast holds. The age-adjustment argument nonetheless rests on the pattern visible in Figure 1C and reproduced in Figure S3C, in which cessation duration rises with age among no-product quitters but is recent at every age among product users, and on the age-matched residual, neither of which depends on a slope model. One quantity is genuinely lost rather than made imprecise: the exposed long-term stratum falls from two respondents to one, so the within-stratum comparison rests on the short-term stratum alone, and the long-term ordering is read from the later and pooled waves (§6, §7).

**Table S3.** Cessation duration under the full and restricted exposure definitions, KNHANES 2018

| Metric | Full product-using group (Table S2) | Restricted to e-cigarette-item responders | Strict variant |
|---|---|---|---|
| Exposed group, n | 43 | 18 | 18 |
| Comparator group, n | 1,305 | 1,330 | 1,305 |
| *A. Cessation duration, all former smokers (years)* | | | |
| Exposed: median | 0.7 | 0.5 | 0.5 |
| Exposed: mean | 1.2 | 1.1 | 1.1 |

| | | | |
|---|---|---|---|
| Comparator: median | 12.0 | 12.0 | 12.0 |
| Comparator: mean | 14.5 | 14.3 | 14.5 |
| Kolmogorov–Smirnov D | 0.76 | 0.73 | 0.75 |
| Cross-group pairs in which the switcher quit more recently | 92% | 92% | 93% |
| ***B. Within the short-term (<5 yr) stratum*** | | | |
| Exposed: median (n) | 0.6 (41) | 0.5 (17) | 0.5 (17) |
| Comparator: median (n) | 2.0 (311) | 1.5 (335) | 2.0 (311) |
| Kolmogorov–Smirnov D | 0.34 | 0.37 | 0.38 |
| Cross-group pairs | 73% | 73% | 74% |
| Exposed respondents in the long-term (>=5 yr) stratum, n | 2 | 1 | 1 |
| ***C. Age adjustment*** | | | |
| Residual gap after exact age-matching (+/- 2 yr), years | 9.3 | 9.5 | 9.5 |
| Age-matched cross-group pairs | 91% | 90% | 91% |
| Theil-Sen slope, exposed (yr per yr of age) | -0.01 | 0.00 | 0.00 |
| Theil-Sen slope, comparator (yr per yr of age) | +0.30 | +0.30 | +0.30 |
| Theil–Sen slope, exposed and comparator (yr per yr of age; 95% CI) | -0.01 (-0.04 to 0.01) vs +0.30 (0.26 to 0.33); non-overlapping | 0.00 (-0.04 to 0.03) vs +0.30 (0.27 to 0.33); non-overlapping | 0.00 (-0.04 to 0.03) vs +0.30 (0.26 to 0.33); non-overlapping |
| ***D. Ceiling on exposed cessation duration*** | | | |
| Longest cessation among exposed respondents, years | 10.0 | 5.0 | 5.0 |
| Long-term comparators who had quit longer ago than any exposed respondent | 70.0% | 93.1% | 93.1% |

*Full product-using group: former smokers reporting current use of liquid EC or HTP (Table S2). Restricted: former smokers answering the umbrella e-cigarette item, with the 25 HTP-only switchers reassigned to the comparator. Strict variant: the same 18 exposed respondents with the 25 HTP-only switchers excluded from both groups. All comparisons are two-sided and unweighted. The Mann–Whitney P values are less than 0.001 in every cell except for the short-term stratum under the restricted definition, where P = 0.002. The Kolmogorov–Smirnov P values in the short-term stratum are 0.019 (restricted) and 0.012 (strict), reflecting the exact test results for 17 exposed observations; in Panel A, all three values are less than 0.001. Slopes are Theil–Sen estimates with distribution-free 95% confidence intervals; the exposed and comparator intervals do not overlap under any of the three definitions. The corresponding ordinary-least-squares interaction coefficients are -0.36 (robust SE 0.024; pooled-variance SE 0.17, P = 0.03), -0.36 (0.023; 0.23, P = 0.12) and -0.35 (0.023; 0.23, P = 0.13) across the three columns; the pooled-variance standard errors are inflated because the exposed group's dispersion is far smaller than the comparator's (§1).*

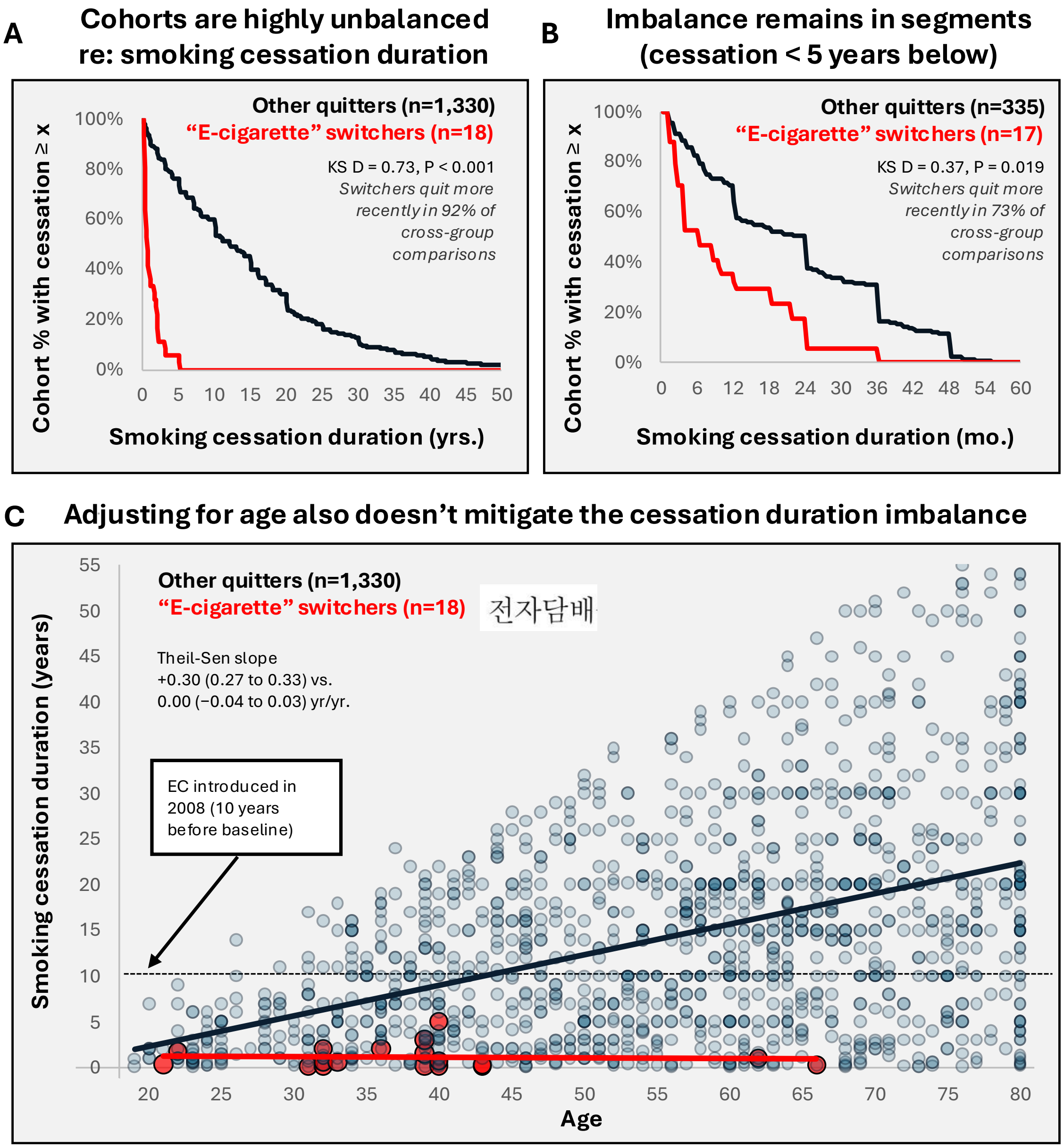


**Figure S3.** Smoking cessation duration under the restricted exposure definition, KNHANES 2018.

*"E-cigarette" switchers (red; n = 18) vs. all other former smokers (blue; n = 1,330). (A) Complementary cumulative distribution across years for all former smokers (Kolmogorov–Smirnov D = 0.73; switchers quit more recently in 92% of cross-group comparisons; P < 0.001). (B) The same in months within the short-term (<5-year) stratum (D = 0.37, 73%, P = 0.019). (C) Age vs. cessation duration with per-group Theil–Sen fits and distribution-free 95% confidence intervals: +0.30 years of cessation per year of age (0.27 to 0.33) among all other former smokers and 0.00 (−0.04 to 0.03) among switchers, nonoverlapping. The dashed line indicates cessation beginning in 2008, when e-cigarettes first went on sale in Korea; HTPs reached the market in June 2017.*

### 6. Replication across KNHANES 2018 to 2023, including age matching

Table S4 repeats the 2018 analyses in each wave from 2018 to 2023, using the year-specific product definitions in §1; the six waves contain 617 product-using and 7,440 no-product former smokers. Because the 2018 product items differed from those used from 2019 (§1), pooled figures combine only the 2019 to 2023 waves (574 and 6,135). In every wave, product users had quit far more recently than other former smokers (median 0.7 to 3.0 years against 12.0 to 15.0; Kolmogorov–Smirnov D = 0.63 to 0.76; product users had quit more recently in 86% to 92% of cross-group pairs), and HTP users made up 66% to 79% of product users.

The two cessation strata behave differently. Within the long-term stratum, the gap persisted in every wave (medians 5.0 to 7.5 years among product users against 16.0 to 19.0 among no-product quitters). Within the short-term stratum, it did not: the difference was 1.4 years in 2018 and 0.5 years in 2019, and from 2020 onward the medians were equal or reversed (2.0 vs 1.6 years in 2023), with no significant separation. The cessation-duration imbalance relevant to Kim et al. is therefore concentrated in the long-term stratum, the stratum that carries their headline estimate (§7).

Exact age matching (±2 years) was repeated in every wave. The matched pool retained 914 to 1,136 no-product former smokers per wave, and the residual cessation gap was 9.3, 9.0, 9.0, 11.0, 11.0 and 10.0 years in 2018 to 2023 (Table S4, Panel E). Cessation duration rose with age among no-product quitters (Theil–Sen 0.29 to 0.38 years per year of age) but not among product users (−0.01 to +0.07), and the confidence intervals did not overlap in any wave. Age is therefore an incomplete proxy for cessation duration in every year examined, which is the basis for the main-text statement that age adjustment cannot remove the imbalance.

Restricting the exposed group to liquid e-cigarette users, possible from 2019 when the e-cigarette item specified liquid products, gives Kolmogorov–Smirnov D = 0.64, 0.59, 0.59, 0.67 and 0.68 in 2019 to 2023 (n = 37 to 46 per wave), against all other former smokers.

**Table S4.** Cessation duration among former smokers by product use, KNHANES 2018 to 2023

| Metric | 2018 | 2019 | 2020 | 2021 | 2022 | 2023 | Pooled 2019–2023 |
|---|---|---|---|---|---|---|---|
| ***A. Sample and product mix*** | | | | | | | |
| Product users, n | 43 | 101 | 98 | 113 | 131 | 131 | 574 |
| No-product former smokers, n | 1,305 | 1,326 | 1,248 | 1,228 | 1,138 | 1,195 | 6,135 |
| Any HTP among product users, % | 79 | 75 | 70 | 66 | 78 | 72 | 72 |
| ***B. All former smokers: years since quitting*** | | | | | | | |
| Median, product users | 0.7 | 2.0 | 2.0 | 3.0 | 2.3 | 3.0 | 2.0 |
| Median, no product | 12.0 | 12.0 | 15.0 | 15.0 | 15.0 | 15.0 | 15.0 |
| Kolmogorov–Smirnov D | 0.76 | 0.68 | 0.67 | 0.68 | 0.69 | 0.63 | 0.65 |
| Product user quit more recently, % of pairs | 92 | 89 | 86 | 86 | 88 | 86 | — |
| ***C. Short-term stratum (<5 years): median years (n)*** | | | | | | | |
| Product users | 0.6 (41) | 1.5 (92) | 2.0 (79) | 2.0 (87) | 2.0 (100) | 2.0 (81) | 2.0 (439) |
| No product | 2.0 (311) | 2.0 (312) | 2.0 (269) | 2.0 (224) | 2.0 (190) | 1.6 (198) | 2.0 (1,193) |

| Metric | 2018 | 2019 | 2020 | 2021 | 2022 | 2023 | Pooled 2019–2023 |
|---|---|---|---|---|---|---|---|
| D (P) | 0.34 (<0.001) | 0.26 (<0.001) | 0.09 (0.67) | 0.12 (0.28) | 0.08 (0.72) | 0.12 (0.32) | 0.07 (0.06) |
| ***D. Long-term stratum (≥5 years): median years (n)*** | | | | | | | |
| Product users | 7.5 (2) | 6.0 (9) | 5.0 (19) | 6.0 (26) | 5.2 (31) | 6.0 (50) | 6.0 (135) |
| No product | 16.0 (994) | 16.0 (1,014) | 18.0 (979) | 19.0 (1,004) | 19.0 (948) | 19.0 (997) | 18.0 (4,942) |
| D (P) | 0.70 (0.18) | 0.80 (<0.001) | 0.68 (<0.001) | 0.69 (<0.001) | 0.66 (<0.001) | 0.70 (<0.001) | 0.66 (<0.001) |
| ***E. Age*** | | | | | | | |
| Median age, product / no product | 39 / 58 | 40 / 58 | 38.5 / 58 | 40 / 60 | 39 / 61 | 39 / 61 | — |
| Age-matched no-product, n | 941 | 1,136 | 955 | 1,068 | 914 | 940 | — |
| Residual gap after age matching, years | 9.3 | 9.0 | 9.0 | 11.0 | 11.0 | 10.0 | — |
| Theil–Sen slope, product users | −0.01 | 0.00 | +0.07 | +0.03 | +0.04 | +0.06 | — |
| 95% CI | −0.04, +0.01 | −0.03, 0.00 | +0.03, +0.11 | 0.00, +0.07 | 0.00, +0.07 | 0.00, +0.12 | — |
| Theil–Sen slope, no product | +0.30 | +0.29 | +0.36 | +0.36 | +0.33 | +0.38 | — |
| 95% CI | +0.26, +0.33 | +0.25, +0.32 | +0.32, +0.40 | +0.32, +0.40 | +0.30, +0.38 | +0.33, +0.42 | — |

*Product users = former smokers currently using HTP or liquid EC (2018: umbrella e-cigarette item or HTP checklist; 2019 onward: liquid EC item, or daily or occasional HTP use). No product = former smokers using neither. Analytic sample: adults aged 19 years or older, valid cessation duration, at least 30 days of abstinence; unweighted. Short-term and long-term strata are defined at 5 years since quitting. P values are two-sided Kolmogorov–Smirnov tests. The 2018 long-term product cell contains two respondents. Age matching retains no-product former smokers within 2 years of age of at least one product user; the residual is the matched median minus the product-user median. Theil–Sen slopes are years of cessation per year of age with distribution-free 95% confidence intervals; the intervals do not overlap in any wave. The pooled column combines the 2019 to 2023 waves, which used the same product items, without weighting; 2018 is excluded from it because its items differed, and respondents differ between waves. Ceiling: the longest abstinence among product users was 10.0 and 9.0 years in 2018 and 2019 and 12.0 to 20.0 years from 2020 onward, so the ceiling described in §4 applies to the baseline years.*

### 7. Published cessation-duration gradient and the long-term stratum

Cessation duration is unmeasured in the NHIS analysis, and it is a strong lung cancer risk factor in this population. In a National Health Insurance Service cohort of 2,974,820 Korean adults, multivariable-adjusted lung cancer hazard ratios among men, relative to continuing smokers, were 0.98 (95% CI 0.91-1.06) below ten years since quitting, 0.53 (0.48-0.58) at ten to under fifteen years, and 0.23 (0.19-0.27) at fifteen years or more [5]. These estimates are adjusted for age and pack-years, so a gradient of this kind is not one that Kim et al.'s covariate set would absorb. Two features limit the transfer: they were computed in men only, because female cases were too few, whereas Kim et al. analyzed both sexes together; and a sensitivity analysis in the same report, restricted to participants already quit at baseline and unable to adjust for pack-years, gives a flatter gradient, 0.57 below ten years falling to 0.35 beyond twenty. We do not attempt to quantify how much of the reported association this gradient would carry. The point is that the variable on which the two groups barely overlap (§4) is one that matters for the outcome, and that it was imprecisely estimated.

Park et al. report three further features relevant here. For all cancers, risk among men who had quit less than ten years earlier was higher than that of continuing smokers (aHR 1.12, 95% CI 1.08–1.16), and the spline for lung cancer rises slightly above 1.0 in the first years after quitting before falling (their Figure 1B). That early excess is absent from their sensitivity analysis based on self-reported cessation duration, which shows a steady decline from quitting, so it may partly reflect the construction of the time-updated categories. Cessation duration was measured from the start of quitting with periods of relapse excluded, and relapsed and transient quitters were retained as separate categories rather than removed (lung cancer aHR 0.90 and 0.72 respectively, against 0.58 for complete quitters) [5].

In patients with COPD, for whom quitting and continued smoking followed the same diagnosis, quitting was associated with lower lung cancer risk within a median follow-up of about five years (aHR 0.85, 95% CI 0.72–0.99) [11]. The early plateau in the general population may therefore partly reflect the circumstances of quitting rather than a delay in biological benefit. Either way, risk among recent quitters is close to that of continuing smokers, and HTP/EC switchers are almost all recent quitters.

Applied to the KNHANES distributions, the relevant contrast is in the long-term stratum. Pooled across 2019 to 2023, long-term product users had quit a median of 6.0 years earlier (n = 135) and long-term no-product quitters 18.0 years earlier (n = 4,942; Kolmogorov–Smirnov D = 0.66, P < 0.001). These durations fall in Park et al.'s under-10-year and 15-or-more-year categories, where lung cancer risk relative to continuing smokers was 0.98 and 0.23. In the short-term stratum, pooled medians were identical (2.0 vs 2.0 years; n = 439 vs 1,193; D = 0.07), within the period in which lung cancer risk changed little after quitting; at that stage the more relevant difference between groups is the likelihood of relapse, which Kim et al. did not track.

### 8. Kim et al.'s high-risk subgroup

Kim et al.'s high-risk subgroup, adults aged 50 to 80 years with at least 20 pack-years, produced their largest incidence estimate for former smokers using e-cigarettes (aHR 1.91, 95% CI 1.44–2.53) [1]. Table S5 applies the same criteria to KNHANES. Pack-years were calculated as cigarettes per day when smoking (BS6_3) divided by 20, multiplied by years smoked (BS6_2 divided by 12). In 2018, 3 product users met both criteria; they had quit 0.25, 1 and 2 years earlier, against a median of 11.0 years among 421 no-product former smokers with nearly identical mean pack-years (37.7 vs 37.6); 86% of the latter had quit before the most abstinent product user.

Pooling the 2019 to 2023 waves, 63 product users in the subgroup had quit a median of 2.0 years earlier (IQR 1.3–5.0) against 12.0 years (6.0–20.0) among 1,906 no-product former smokers (Kolmogorov–Smirnov D = 0.60, P < 0.001); product users had quit more recently in 86% of cross-group pairs. Mean pack-years were 33.8 and 37.7, so product users in the subgroup had, if anything, slightly lower cumulative exposure, and they were younger (mean age 58.9 vs 67.5 years). The gap was present in every wave. Eligibility criteria select on age and cumulative exposure; they do not balance time since cessation, and the subgroup that produced Kim et al.'s largest estimate shows a cessation gap of about a decade.

**Table S5.** Years since quitting among former smokers aged 50 to 80, with and without the ≥20 pack-year criterion, KNHANES 2018 to 2023

| **Metric** | **2018** | **2019** | **2020** | **2021** | **2022** | **2023** | **Pooled 2019–2023** |
|---|---|---|---|---|---|---|---|
| ***A. Ages 50 to 80 years*** | | | | | | | |
| Product users / no product, n | 4 / 858 | 15 / 861 | 16 / 820 | 26 / 862 | 28 / 795 | 21 / 858 | 106 / 4,196 |
| Median years since quitting (IQR), product users | 0.6 (0.3–1.3) | 1.0 (0.9–1.7) | 3.5 (2.0–5.0) | 3.0 (1.4–5.0) | 2.5 (1.5–4.0) | 4.0 (2.0–6.0) | 2.3 (1.0–5.0) |
| Median years since quitting (IQR), no product | 15.5 (7.0–25.0) | 15.0 (7.0–25.0) | 19.8 (9.0–30.0) | 20.0 (10.0–29.0) | 20.0 (10.0–30.0) | 20.0 (10.0–30.0) | 19.0 (10.0–29.0) |
| Kolmogorov–Smirnov D (P) | 0.90 (<0.001) | 0.81 (<0.001) | 0.71 (<0.001) | 0.74 (<0.001) | 0.72 (<0.001) | 0.71 (<0.001) | 0.71 (<0.001) |
| Product user quit more recently, % of pairs | 95 | 95 | 88 | 91 | 92 | 89 | 90 |
| No product quit before longest-abstinent product user, % | 90 | 78 | 67 | 67 | 59 | 68 | 57 |
| Mean pack-years, product / no product | 32.1 / 22.4 | 28.5 / 22.7 | 24.6 / 21.1 | 26.3 / 21.6 | 25.3 / 20.5 | 21.2 / 20.5 | 25.1 / 21.3 |
| Mean age, product / no product | 62.5 / 65.7 | 59.0 / 65.7 | 57.4 / 66.0 | 58.6 / 66.5 | 56.0 / 66.3 | 57.4 / 66.1 | 57.6 / 66.1 |
| ***B. Ages 50 to 80 years and ≥20 pack-years (Kim et al. high-risk subgroup)*** | | | | | | | |
| Product users / no product, n | 3 / 421 | 9 / 417 | 9 / 364 | 14 / 399 | 19 / 350 | 12 / 376 | 63 / 1,906 |
| Median years since quitting (IQR), product users | 1.0 (0.6–1.5) | 1.0 (0.9–1.2) | 5.0 (2.0–8.0) | 2.0 (1.6–4.5) | 2.0 (1.3–3.2) | 4.0 (2.0–6.5) | 2.0 (1.3–5.0) |
| Median years since quitting (IQR), no product | 11.0 (5.0–20.0) | 10.0 (5.0–19.0) | 12.0 (5.3–20.0) | 14.0 (7.0–20.0) | 12.6 (6.7–20.0) | 13.0 (7.6–20.0) | 12.0 (6.0–20.0) |
| Kolmogorov–Smirnov D (P) | 0.86 (0.006) | 0.82 (<0.001) | 0.52 (0.01) | 0.73 (<0.001) | 0.74 (<0.001) | 0.54 (<0.001) | 0.60 (<0.001) |
| Product user quit more recently, % of pairs | 92 | 93 | 76 | 89 | 91 | 81 | 86 |
| No product quit before longest-abstinent product user, % | 86 | 67 | 48 | 73 | 56 | 52 | 50 |
| Mean pack-years, product / no product | 37.7 / 37.6 | 37.8 / 38.8 | 33.3 / 38.3 | 37.9 / 38.0 | 32.4 / 37.1 | 28.9 / 36.2 | 33.8 / 37.7 |
| Mean age, product / no product | 66.3 / 67.0 | 60.4 / 66.8 | 57.2 / 67.1 | 62.4 / 67.9 | 56.1 / 68.2 | 59.2 / 67.5 | 58.9 / 67.5 |

*Definitions of product use, analytic sample and tests as in Table S4. Pack-years = (cigarettes per day ÷ 20) × years smoked; respondents with missing smoking duration or intensity are excluded from Panel B. IQR, interquartile range. Cells with fewer than ten product users (2018 and 2019 to 2020 in Panel B, and 2018 in Panel A) are descriptive only. The pooled column combines the 2019 to 2023 waves without weighting; 2018 is excluded from it because its product items differed.*

#### *Cessation duration of high-risk long-term quitters*

Kim et al. reported no cessation duration for any group, so the duration of their high-risk long-term quitters, the comparator carrying the strongest subgroup claims, cannot be read from the article. It can be estimated by mapping that group onto KNHANES respondents matched on the covariates the article does report. Restricting KNHANES 2018 to no-product former smokers aged 56 to 71 (the mean plus or minus one standard deviation of Kim et al.'s long-term high-risk quitters without e-cigarette use; Extended Data Table 2), with at least 20 pack-years, who had quit at least 5 years before baseline (the article's long-term definition), yields a group matched on age and smoking intensity (Table S6).

The covariate match is close: the mean age is 64.0 years, whereas it is 63.6 years, and the mean number of pack-years is 33.8 and 30.4, respectively. The median cessation duration was 15.0 years (IQR 10.0 to 20.0), and 77% had quit at least 10 years before baseline. In KNHANES 2018, no product user had quit more than 10 years before the survey (Table S2), consistent with the arrival of e-cigarettes in Korea in 2008. On this mapping, about two-thirds (67%) of the high-risk comparison group had quit longer ago than any product user in KNHANES 2018. This is an inference from an external survey rather than a measurement of the NHIS cohort; the two are national and contemporaneous but not identical, pack-years are reconstructed from self-reported duration and daily amount, and the matched cell contains 173 respondents.

**Table S6.** Cessation duration of KNHANES respondents matched to Kim et al.'s high-risk long-term quitters

| Metric | KNHANES analog | Kim et al. group |
|---|---|---|
| Mean age (yr) | 64.0 | 63.6 |
| Mean pack-years | 33.8 | 30.4 |
| Median cessation (yr) | 15.0 (IQR 10.0-20.0) | not reported |
| Quit >=10 years | 77% | — |
| Quit >=15 years | 53% | — |
| Quit >=20 years | 31% | — |
| n | 173 | 340,655 (long-term, no e-cigarette) |

KNHANES analog: no-product former smokers aged 56 to 71 years, at least 20 pack-years, who quit at least 5 years; KNHANES 2018 (n = 173). Kim et al. group: high-risk (50 to 80, at least 20 pack-years) long-term quitters without e-cigarette use, Extended Data Table 2 (n = 340,655); the article reports mean age and pack-years but no cessation duration. Cessation figures are estimated from the matched KNHANES respondents and are not measured in the NHIS cohort.

### 9. Age adjustment acting on sparse events

The cessation asymmetry documented in §4 also appears in Kim et al.'s own person-time, which is why the model adjusts the exposed cell using participants with whom the exposed cells barely overlap. Among HTP/EC-exposed participants, 27,046 of 136,459 person-years (19.8%) fall in the long-term cessation stratum; among quitters using no product, 7.56 of 11.64 million (64.9%) do. Within that stratum, the exposed contributes one person-year for every 279 in the reference stratum and one lung cancer event for every 714 (Kim et al., Table 2). The two groups therefore occupy opposite ends of the cessation–duration axis, and the covariate structure adjusting the 16 exposed events is drawn from a distribution that those events do not share.

This is the adjustment the main text describes. In the long-term stratum, the crude lung cancer incidence among HTP/EC switchers is 5.9 per 10,000 person-years, whereas it is 15.1 among no-product quitters (Kim et al., Table 2); thus, the unadjusted rate is 61% lower in the exposed group. Full adjustment (model 2) converts this into an adjusted hazard ratio of 1.74 (95% CI 1.07 to 2.84), a reversal of direction and a 4.5-fold change from the crude estimate. The exposed cells contribute 16 lung cancer events, compared with 11,420 events in the reference. Sixteen events against ten covariates is approximately 1.6 events per covariate, well below the ten events per covariate conventionally treated as a minimum for stable Cox estimation; an adjustment of this size therefore rests on a cell too sparse to support it, drawn from a covariate distribution that the exposed does not share.

The concern that age adjustment can amplify rather than correct an estimate built on a handful of events is not hypothetical. Plurphanswat, Selya and Rodu [14] reanalyzed the National Health Interview Survey (2014 to 2021) data underlying a widely cited report associating e-cigarette use with myocardial infarction. Among never smokers who currently used e-cigarettes, 12 myocardial infarction events were observed. The crude odds ratio was 0.42 (95% CI 0.24–0.75); adjustment for every covariate except age moved it to 0.80 (0.45–1.43), still below 1; adding age to that model reversed it to 2.48 (1.35–4.55), and age added alone to the crude model gave 3.00 (1.66–5.43). The reversal was therefore produced by age acting on 12 exposed events whose ages sat in the upper tail of an otherwise young exposed group.

The structural parallel to that of Kim et al. is close: a protective crude estimate, an exposed cell of comparable size (16 lung cancer events in the long-term stratum), an exposed group approximately a decade younger than its reference, and an adjustment in which a minimal model (age, sex and pack-years) produces nearly all of the shift in the pooled estimate (Model 1 versus Model 2, 1.58 versus 1.56). Whether the same amplification occurs here cannot be determined from the published tables because the age distribution of cases within each exposure group is not reported. We therefore note the possibility rather than assert it and identify the reporting that would resolve it: case age distributions by exposure group or an analysis stratified on age within the long-term stratum.

### 10. Exposure classified once: time-varying analysis

The exposure in the primary analysis is fixed at the 2018 screening. A participant who was a dual user at baseline and switched completely thereafter therefore remained in the current-smoker comparator for the whole follow-up period. Table S1 is consistent with movement in that direction: the dual-user to switcher ratio fell from 5.2:1 in 2018 to 2.1:1 in 2019. These are independent cross-sections, and the 2019 items were product-specific, so the change cannot be read as individual transitions; to the extent that dual users went on to switch, however, the comparator accumulated the exposure under test.

#### Time-varying exposure analysis

Kim et al. address this with time-varying models in which smoking status and e-cigarette use are treated as time-dependent covariates updated at the follow-up screening date. The analysis is restricted to the subpopulation rescreened from 2019 to 2020 (n = 3,465,275 of 4,524,895, or 76.6%), and the authors describe this as an exploratory subpopulation. Table S7 presents every estimate from that analysis beside its fixed-exposure counterpart, on the common contrast used in the main text: e-cigarette-using ex-smokers against ex-smokers not using a product, fully adjusted (model 2). All six estimates are lower under time-varying exposure than under fixed exposure. The direction of that shift is what misclassification of the comparator would produce, but the magnitude cannot be attributed to reclassification alone because the rescreened subpopulation is smaller and the event counts decrease correspondingly. Supplementary Tables 7 and 8 report only event counts and hazard ratios: no group sizes, no person-years, and no count of participants who changed exposure category. Without those, reclassification and reduced power cannot be separated, and we therefore report the pattern without attributing it to a single cause.

**Table S7.** Fixed-exposure and time-varying estimates compared e-cigarette-using ex-smokers vs. ex-smokers who did not use a product (Kim et al., Model 2)

| Outcome | Fixed: events | Fixed: aHR (95% CI) | Time-varying: events | Time-varying: aHR (95% CI) |
|---|---|---|---|---|
| ***Overall cohort*** | | | | |
| Lung cancer incidence | 71 | 1.56 (1.24-1.97) | 31 | 1.38 (0.98-1.98) |
| Lung cancer-specific death | 19 | 2.00 (1.28-3.15) | 3 | 0.77 (0.25-2.39) |
| All-cause mortality | 170 | 1.22 (1.05-1.41) | 75 | 1.18 (0.94-1.48) |
| ***High-risk subgroup (50-80 yr, >=20 pack-years)*** | | | | |
| Lung cancer incidence | 49 | 1.91 (1.44-2.53) | 14 | 1.40 (0.83-2.37) |
| Lung cancer-specific death | 14 | 1.92 (1.13-3.24) | 3 | 1.16 (0.37-3.61) |
| All-cause mortality | 62 | 0.97 (0.76-1.25) | 18 | 0.79 (0.50-1.25) |

*Fixed-exposure estimates from Kim* et al.*, Extended Data Tables 1 and 3; time-varying estimates from Supplementary Tables 7 and 8. All are model 2 (adjusted for age, sex, pack-years, body mass index, household income, alcohol consumption, physical activity, Charlson comorbidity index, COPD and idiopathic pulmonary fibrosis), with ex-smokers not using a product as the reference. Note that the reference group key printed beneath Extended Data Tables 1 and 3 and Supplementary Tables 7 and 8 describes short-term and long-term cessation strata, which those four tables do not contain; the reference groups above are read from the rows marked as references in each table.*

Cessation duration is not a covariate in any model; it enters only as a level of the exposure classification in the five- and six-group analyses, and is absent from the three-group classification used in every time-varying table. The same reference-key mismatch noted beneath Table S7 applies to Supplementary Tables 17 and 18, whose estimates are likewise against all product-free ex-smokers pooled.

#### What pooling does to the reference group

Because the time-varying analyses pool the cessation strata, the hazard of their reference group is set by its long-term majority. This information can be read directly from the article. Supplementary Tables 10 and 11 report the same cohort under the same any-use exposure definition, pooled and stratified, respectively, and their event counts are exactly reconciled; thus, pooling is the only difference between them. Among product-free ex-smokers, the short-term stratum contributes 6,971 lung cancer events at an adjusted hazard of 0.66 relative to current smokers, and the long-term stratum contributes 11,399 events at 0.51; the pooled value is 0.56, which is the event-weighted geometric mean of the two (0.562). The exposed arm blends in the opposite direction, with 115 short-term events at 0.96 and 37 long-term events at 0.63, pooling to the reported 0.85. The pooled contrast, 0.85 divided by 0.56, reproduces the reported 1.52 and is therefore a comparison between an exposed group that is 76% short-term by events and a reference group that is 62% long-term. Updating exposure status over follow-up does not alter that composition, and no covariate in the model represents it.

### 11. Precision, product mix and exposed events

For every estimate in Kim et al., the standard error of the log hazard ratio recovered from the reported confidence interval matches 1/√(exposed events) to within 0.002 (Table S8). Precision is therefore set by the number of exposed events; the reference group's thousands of events add almost nothing, and the size of the cohort does not translate into precision for the exposure under study.

If a fraction p of the exposed group used liquid e-cigarettes, the observed log hazard ratio approximates a p-weighted mixture of the liquid e-cigarette and HTP effects. Recovering the e-cigarette effect divides by p, so its standard error scales by 1/p and the number of exposed events needed for the same precision by $1/p^2$. In KNHANES 2018, 9 of 18 product-using former smokers who affirmed the umbrella item reported no HTP use (p = 0.50), and 9 of 43 product users overall (p = 0.21), giving multipliers of 4 and about 23. The calculation assumes the HTP effect is known; without that assumption the e-cigarette effect is not identified. The final two columns of Table S8 express each estimate as the number of events an e-cigarette-specific estimate would effectively rest on.

**Table S8.** Exposed events and precision of Kim et al.'s estimates

| Estimate | Exposed events | SE from reported CI | 1/√events | EC-equivalent events, p = 0.50 | EC-equivalent events, p = 0.21 |
|---|---|---|---|---|---|
| Pooled incidence | 71 | 0.118 | 0.119 | 17.8 | 3.1 |
| Long-term stratum incidence | 16 | 0.249 | 0.250 | 4.0 | 0.7 |
| Pooled lung cancer death | 19 | 0.230 | 0.229 | 4.8 | 0.8 |
| Time-varying incidence | 31 | 0.179 | 0.180 | 7.8 | 1.4 |
| Time-varying lung cancer death | 3 | 0.576 | 0.577 | 0.8 | 0.1 |
| Dual users vs cigarette-only | 578 | 0.042 | 0.042 | 67.6 (p = 0.34) | 31.7 (p = 0.23) |

*SE = ln(upper/lower)/3.92 for the log hazard ratio. EC-equivalent events = exposed events × $p^2$. For dual users, p is taken from KNHANES 2018 current smokers: 52 of 152 umbrella-item endorsers (0.34) and 52 of 222 product users (0.23). Estimates from Kim et al., Table 2, Extended Data Table 1 and Supplementary Tables 1 and 7.*

The comparison in Kim et al. least affected by cessation timing is dual users against cigarette-only smokers (their Supplementary Tables 1 and 2): aHR 0.78 (0.72–0.85) on 578 events overall and 0.94 (0.83–1.07) on 252 events in the high-risk subgroup [1]. Both groups were current smokers, so cessation timing could not differ between them. Product mix applies here as well: in KNHANES 2018, 77% of product-using current smokers reported HTP use (170 of 222). Kim et al. report no cigarettes-per-day figures for dual users, so reduced smoking intensity cannot be separated from any product effect. In the authors' 2024 conference abstract, dual users formed a separate exposure group; in the published paper they are part of the current-smoker reference. We report this estimate as evidence about design, not about products.

### 12. Design of NHIS-based studies of cessation and product use

The main text proposes that the limitations of Kim et al. can be addressed within the same national database. Table S9 compares three analyses of NHIS screening data: Kim et al., Kim T. et al. in patients with COPD, and Park et al. on cancer risk after smoking cessation. They differ chiefly in how often smoking and product use were assessed and in what was done when behavior changed.

**Table S9.** Design of three NHIS-based analyses of smoking cessation, product use and lung cancer

| Feature | Kim et al. [1] | Kim T. et al. [11] | Park et al. [5] |
|---|---|---|---|
| Population | 4,524,895 adults with a smoking history, screened in 2018 | 23,262 adults aged 40 to 80 who smoked at COPD diagnosis, 2016 to 2023 | 2,974,820 adults aged 30 or older; time-since-quitting analyses in 1,727,340 men |
| Smoking assessments used | Two: 2012 to 2014 and 2018; one further update in 2019 to 2020 for 76.6% (time-varying models) | Two: before (Exam 1) and within 3 years after diagnosis (Exam 2) | Every biennial screening, 2002 to 2017 (up to 8 cycles) |
| Product exposure | Single umbrella e-cigarette item (2018); daily use | Any current e-cigarette use at Exam 2 (1,316 users, of whom 923 dual users) | None (cigarettes only) |
| Time since quitting | Two strata (quit after vs before 2012 to 2014); not modeled | Balanced by design: quitting and switching occurred between the same two examinations | From start of quitting, relapse periods excluded; <10, 10 to <15, ≥15 years |
| Relapse | Not tracked, except in the time-varying subsample | Sensitivity analysis restricted to patients maintaining their category | Relapsed and transient quitters kept as separate categories |
| Comparator | Quitters not using e-cigarettes | Continuing smokers | Continuing smokers |
| Lung cancer estimate | Switchers vs quitters: 1.56 (1.24–1.97), 71 events; long-term 1.74 (1.07–2.84), 16 events; time-varying 1.38 (0.98–1.98), 31 events | Switchers 0.94 (0.65–1.37), 30 events; quitters 0.85 (0.72–0.99) | Men who quit <10 years 0.98 (0.91–1.06); 10 to <15 years 0.53 (0.48–0.58); ≥15 years 0.23 (0.19–0.27) |

*Hazard ratios are adjusted (95% CI). Kim et al.: Table 2, Extended Data Table 1 and Supplementary Table 7. Kim T. et al.: Table 2; switchers include dual users. Park et al.: Tables 2 and 3. The three studies draw on the same national screening database and may share participants; they are compared for design, not pooled.*

Kim T. et al. avoided the cessation-timing imbalance by comparing people whose behavior changed between the same two examinations, and found no lung cancer excess among switchers relative to continuing smokers. When they restricted the analysis to patients who maintained their reported category at later screenings, 24% of switchers were retained against 68% of continuing smokers (their Supplementary Table 5), so persistence of switching could be examined only in a small subset. Park et al. showed that biennial reassessment over up to eight cycles is feasible in the same database and keeps relapsers in the analysis as categories rather than excluding them. Applied to product use, with the separate HTP and liquid e-cigarette items available from 2019, that approach would retain all exposed events and allow dose-response estimates by duration of use for each product.
